# Release-aware, SAS-equivalent Elixhauser comorbidity scoring in R: cross-implementation validation and application to Texas inpatient discharges

**DOI:** 10.64898/2026.10.01.26364551

**Authors:** Minh Tran, Dong Pei

## Abstract

The Elixhauser comorbidity measures are among the most widely used risk adjusters in administrative health data. Their reference implementation, published by the Agency for Healthcare Research and Quality (AHRQ) as SAS programs, has eleven annual releases in two families, one screening pre-existing conditions on the present-on-admission (POA) indicator and the earlier one on Medicare Severity Diagnosis-Related Group, and no implementation of every release existed outside SAS. We developed ecsr10, an open-source R package covering all eleven releases of both families, with a browser-based application over the same functions, and validated it against AHRQ’s own SAS programs over 40,027,114 value-level comparisons with zero disagreements. Two existing open-source reimplementations scored on the same dataset showed reproducible defects. We then used the package to quantify two choices AHRQ’s software leaves to the analyst and studies seldom report, the release and the handling of diagnoses not present on admission, on the Texas Inpatient Public Use Data File, 2016 Q1–2019 Q4 (8,585,244 adult discharges from 726 hospitals), scoring one predefined cohort under every release and pre-existing-condition setting. POA handling dominated: disabling POA changed the comorbidity profile of 56.7% of admissions, and a POA-naive mortality index scored higher on discrimination (area under the curve 0.807 versus 0.789; difference 0.0183, 95% confidence interval 0.0161–0.0204) by counting in-hospital complications as pre-existing disease. POA reporting was bimodal across hospitals, so pooled hospital comparisons on such a file partly compare documentation practice. Because the cohort predates every release compared, the release contrast is a lower bound: the release chosen between v2022.1 and v2026.1 changed at least one comorbidity flag for 0.0036% of admissions, yet a single revision of ten mortality weights changed the comorbidity index for 42.4% of them. An exactly validated implementation makes such choices measurable; release, POA handling and the implementation used should be reported as study characteristics.

**Author summary:** Hospital discharge records are routinely used to compare hospitals, adjust for how sick patients were on arrival, and predict outcomes. Much of that work rests on comorbidity measures maintained by a United States federal agency, whose reference software exists only in SAS and has changed eleven times. We built a free package that reproduces the agency’s programs exactly in every version, checked it against the agency’s own programs with no disagreement, and found that two existing free reimplementations do not. We then used it to score 8.6 million Texas hospital admissions every way the agency’s software allows, to ask which of the choices studies rarely report change conclusions. Far more consequential than the annual version was whether the software is told which diagnoses patients had on arrival. Ignoring that information counts hospital complications as pre-existing illness. It changed the comorbidity profile of more than half of admissions, and made a mortality model look more accurate while measuring something else. Hospitals also differed sharply in whether they recorded that information. The annual version barely moved the comorbidity flags, yet one revision of the scoring weights changed the risk score for 42% of admissions.

## Introduction

Comorbidity adjustment is a routine component of research on administrative hospital data. The Elixhauser system, introduced in 1998 as 30 binary comorbidity indicators derived from discharge diagnosis codes [1], is one of the two dominant approaches alongside the Charlson index [2] and has been repeatedly translated as coding systems changed [3]. Summary scores derived from it, van Walraven’s point system [4] and AHRQ’s own readmission and mortality indices [5], serve as single-number covariates in outcome models, hospital profiling and health services research, and perform at least as well as competing indices for inpatient mortality [6–10].

The Agency for Healthcare Research and Quality (AHRQ), through the Healthcare Cost and Utilization Project (HCUP), maintains the reference implementation as a set of SAS programs [11]. Three features of that reference are consequential and easy to overlook.

First, the reference is not a single program. AHRQ has published eleven releases since October 2015 in two structurally distinct families. The *beta* family (v2016.2 through v2020.1) assigns 30 measures and excludes secondary diagnoses that fall inside 24 Medicare Severity Diagnosis-Related Group (MS-DRG) screens, because a comorbidity must be distinguishable from the reason for admission; it therefore applies to inpatient data only. The *Refined* family (v2021.1 through v2026.1) replaces the MS-DRG screen with the present-on-admission (POA) indicator, expands to 38 measures, and from v2022.1 adds two published index scores. The two families read different columns, emit different columns, and answer different questions about the same admission.

Second, each release is tied to a diagnosis-code version. For the 18 measures that depend on presenton-admission status, a diagnosis on the annual POA-exempt list of the International Classification of Diseases, Tenth Revision, Clinical Modification (ICD-10-CM) is treated as pre-existing whatever was reported. That list is version-specific, and the SAS program derives the version from the discharge date through a fiscal-year mapping that each release truncates at its own maximum, so the same admission can score differently under two releases for a reason that is indistinguishable from ordinary code-list drift unless releases are compared on the same dates.

Third, present-on-admission information is optional in practice. POA reporting became a Medicare requirement for inpatient claims under the Deficit Reduction Act of 2005, but completeness varies widely across data sources and hospitals. That diagnosis-timing information changes measured comorbidity, safety indicators and case mix is well established in general [12–14]; what has not been measured is how much it changes the output of AHRQ’s specific software, under its specific POA conventions and releases, on a population-scale file. AHRQ’s software offers a switch for data without POA, and analysts faced with such a file choose ad hoc among several plausible alternatives.

These features create a reproducibility problem that is specific and testable. An analyst reporting “Elixhauser comorbidities, AHRQ software” has not specified the analysis: the statement is compatible with eleven code tables, two incompatible screening mechanisms, three or more POA conventions and, before v2023.1, a different set of mortality weights. Reporting guidance for routinely collected health data asks for exactly this level of algorithm specification [15], and it is rarely supplied. Meanwhile, analysts working outside SAS depend on reimplementations. A widely used general-purpose R package [16] implements the classic Elixhauser and Charlson mappings rather than AHRQ’s Refined releases, and the two packages that do target them, the R package medicalcoder [17] and the Python package risk-compose [18], have not, to our knowledge, been validated cell by cell against AHRQ’s programs. Software error is a recognized and under-measured contributor to irreproducible computational research [19–21], and a wrong comorbidity flag produces a plausible number, not an error message.

This study makes two contributions, and the first is what makes the second possible. The first is ecsr10, an open-source R package implementing both families and all eleven releases of AHRQ’s software, with a browser-based application over the same functions for users who do not write analysis code, together with its validation against AHRQ’s own SAS programs run live, over three reference datasets and 40,027,114 value-level comparisons, and with the two independent reimplementations we could identify scored on the same dataset and adjudicated against the SAS output. Any measurement of what the software’s options do is only as trustworthy as the implementation that scores them, and none covering every release existed outside SAS.

The second is an applied study using that package to ask which of the choices AHRQ’s software leaves to the analyst actually change a result, and by how much. We scored one predefined cohort from an all-payer state discharge census of 8,585,244 admissions under every release and every pre-existing-condition convention the software supports, and measured the effect of each choice on comorbidity prevalence, the two index scores, out-of-sample mortality discrimination and hospital-level case mix. We expected, before scoring, that release differences would be smaller than POA-handling differences in this cohort, because the Texas files end in 2019 and so predate every release compared, which leaves only code-table content able to contribute to a release difference (see Methods). The study was designed to test that expectation and measure the size of the difference, not to select, from fourteen scoring configurations, whichever showed the largest change.

## Materials and methods

### Software, interfaces, and availability

ecsr10 is an R package, distributed at https://github.com/dpei/ecsr10, that implements both families of AHRQ’s Elixhauser Comorbidity Software for ICD-10-CM behind two entry points. comorbidity() takes a wide data frame, the names of the diagnosis columns, and either the POA columns with year and quarter columns (Refined family) or the MS-DRG column (beta family), and returns the input with the comorbidity flags bound on. cmr_index() adds the two index scores. One argument selects the family and another selects the release; both default to Refined v2026.1, the most recent release.

Three design decisions are relevant to the validation that follows. The first is that release-awareness is carried by data rather than by code: each release’s code table is a slice of one packaged dataset parsed directly from AHRQ’s SAS format programs, and the POA-exempt lists are keyed by ICD-10-CM version rather than by release, because every release ships identical lists for the versions it shares with its predecessor, a property we verify during data generation rather than assume. The second is that the date-to-version mapping is truncated at each release’s own maximum: determine_icd_version() maps discharge year and quarter to ICD-10-CM version 33–43 following AHRQ’s fiscal-year rollover, then caps the result at the maximum version the selected release ships, and dates outside AHRQ’s mapping default to that cap, as they do in the SAS program.

The third is that scoring operates on sets of codes rather than on one encounter at a time. The diagnosis columns are stacked into a long table with one row per diagnosis code, each release’s lookup patterns are matched once against the distinct code vector, and overlapping matches are resolved by the lookup table’s own row order (Fig 1). Optional multicore execution splits rows into contiguous blocks, with all lookup tables built before forking, so that per-block results are identical to a whole-dataset run.

**Fig 1.**
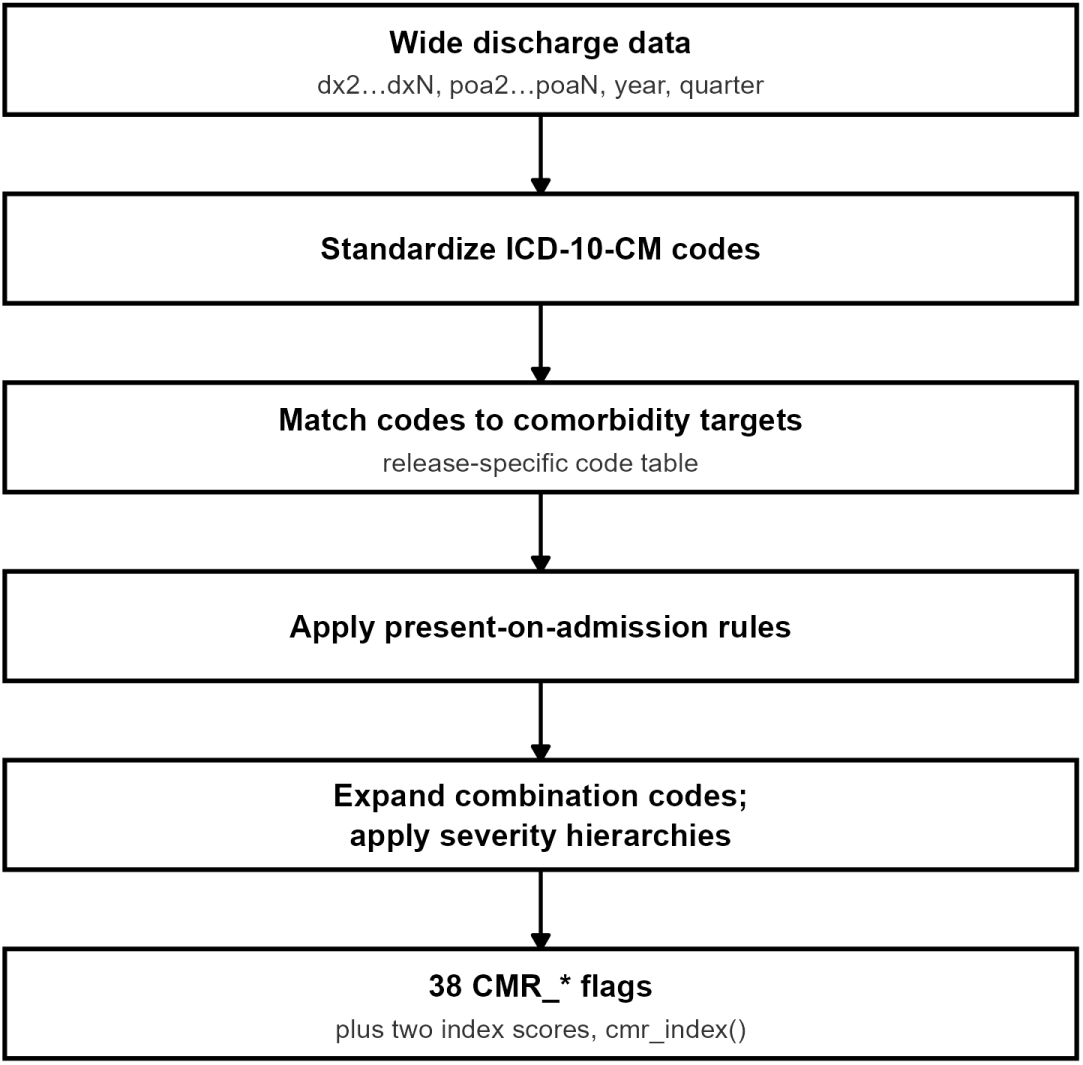
Schematic of the ecsr10 scoring workflow from wide diagnosis data to comorbidity flags. `comorbidity()` reshapes the diagnosis columns, matches each distinct code against the selected release’s code table, applies the present-on-admission rules, expands the combination codes and applies the severity hierarchies, and returns the input with the comorbidity flags bound on. `cmr_index()` adds the two index scores. The beta family follows the same path with its MS-DRG exclusion screen in place of the present-on-admission step, and publishes no indices.

When POA is unavailable, comorbidity() can be run with POA scoring disabled, which reproduces AHRQ’s POA-off macro setting exactly: the 18 POA-dependent measures are returned as missing, not zero, because AHRQ’s program initializes them only inside its POA branch. The package offers no mode that scores those measures without POA data; an analyst who needs all 38 measures from a file without POA has to state that assumption in the data, by setting every POA value to Y, and score with POA enabled.

The package is complemented by a browser-based application, available at https://slicer.shinyapps.io/ecsr10/, for users who prefer not to write analysis code. Users upload a wide-format file, choose a family and release, review a per-column summary naming the role each column plays, and download encounter-level flags with the two indices. The interface scores nothing itself: it is a wrapper over the exported comorbidity() and cmr_index(), so the browser path and the R path run the same code. Uploads are held in memory for the life of the session and are not persisted. The application states that it is a demonstration tool in an environment not compliant with the Health Insurance Portability and Accountability Act (HIPAA), and it keeps scoring disabled until the user confirms the file carries no protected health information.

### AHRQ reference implementations

AHRQ’s own SAS programs are the reference against which every result below is validated. Table 1 summarizes the two families. Both take a wide discharge record with a principal diagnosis and up to *n* secondary diagnoses; the methodology applies to secondary diagnoses only, so the principal diagnosis is never scored.

**Table 1.** The two AHRQ Elixhauser Comorbidity Software families implemented by `ecsr10`.

| Property | Refined | Beta |
| --- | --- | --- |
| Releases | v2021.1 – v2026.1 (six) | v2016.2 – v2020.1 (five) |
| ICD-10-CM versions covered | ICD-10-CM v33 – v43 | ICD-10-CM v33 – v37 |
| Comorbidity measures | 38 | 30, plus a derived hypertension union |
| Pre-existing-condition test | present-on-admission, on 18 of 38 measures | 24 MS-DRG exclusion screens |
| Index scores | readmission and mortality, from v2022.1 | none |
| Additional input required | POA indicators; discharge year and quarter | MS-DRG |
| Applicable data | inpatient and, from v2021.1, other settings | inpatient only |

In the Refined family, 20 measures are assigned from the diagnosis code alone and 18 only when the diagnosis was present on admission, meaning it was reported as Y or W or appears on the ICD-10-CM POA-exempt list for the version the discharge date resolves to. Ten combination codes map to multiple measures, six severity hierarchies suppress the milder member of a pair, and cerebrovascular disease is derived from three intermediate states in which an acute code not present on admission overrides an otherwise qualifying sequelae code. In the beta family, statement order in AHRQ’s DATA step is itself part of the specification: the severity hierarchy runs before the MS-DRG screen, and the hypertension union is derived after it.

### Reference-validation design

We generated the reference by running AHRQ’s own unmodified SAS programs on SAS OnDemand for Academics through a scripted driver. One SAS macro setting is worth stating because agreement depends on it. AHRQ’s programs bound their diagnosis loop at the smaller of a per-record diagnosis count and the number of diagnosis columns supplied. From v2022.1 the count can be left blank, so every supplied column is read, and the reference runs used that documented form; v2021.1 and the five beta programs hardcode the bound, so for those the test datasets carry a diagnosis count equal to each record’s true filled count. Under both configurations the SAS reads exactly the codes present, which is what makes a column-for-column comparison with ecsr10 well defined.

Three reference datasets were then scored under every release each covers:

1. **Vendored AHRQ reference dataset**: 167,322 encounters of AHRQ’s own SAS output, vendored with attribution from the test material of the medicalcoder package [17], one of the two implementations evaluated below. The file contents are the output of AHRQ’s program, a federal work product not subject to copyright. Scored under v2022.1–v2026.1, it validates the content of every release’s code table.
2. **Multi-release edge-case test dataset**: 11,954 encounters purpose-built for this study, spanning 67 distinct (year, quarter) cells from 2014 to 2028 plus missing and invalid dates, and containing all 4,773 codes mapped by any Refined release. It is the only dataset on which a single row’s answer depends on the release, and the only one that reaches ICD-10-CM versions 33–38. Labeled strata cover the version sweep, the full cerebrovascular truth table, every combination target and hierarchy, each measure in isolation, diagnosis-position edges and a random sample, and its release-discriminating rows are themselves verified to discriminate. It was scored twice under each release: with POA enabled, and with POA disabled against SAS run with its POA-off macro setting.
3. **Beta MS-DRG test dataset**: 6,018 encounters covering 100% of every beta version’s code list and both endpoints of every screen range plus the DRGs immediately outside them.

Every scoring configuration was compared with the same script, reused verbatim across all validation pipelines with its checksum asserted at run time. The criterion is value-level equality on the shared columns, matched by name and row-aligned on the encounter identifier, at a tolerance of 10-10; byte identity and column order are not part of it, because AHRQ’s output carries a metadata column we do not emit and orders its columns differently.

### Independent implementation comparison

We additionally scored the multi-release test dataset with the two independent reimplementations we could identify that target the same AHRQ releases, the R package medicalcoder 0.9.0 [17] and the Python package risk-compose 1.0.2 [18], each run unmodified in an isolated environment. Because ecsr10 matches the SAS on every compared cell, each divergence these packages show is equally a divergence from AHRQ’s program, so disagreements can be adjudicated against the SAS output rather than merely reported. For medicalcoder we separated a missing capability from a defect by re-running ecsr10 with that package’s own date-blind POA-exempt table. risk-compose does not run to completion on this input, so each of its divergences was measured as the difference between successive patch levels. A small dataset of deliberately non-conformant input was scored by all four implementations to describe, outside the validation criterion, how each treats input AHRQ documents as invalid. Design detail is in S1 Text.

### Texas data source and cohort

We used the Texas Inpatient Public Use Data File (PUDF) for 2016 Q1 through 2019 Q4, released by the Texas Department of State Health Services through the Texas Health Care Information Collection (THCIC) program [22]. The file is an all-payer census of non-federal Texas hospital discharges. It reports up to 24 secondary diagnoses, each paired with its own POA indicator, along with the discharge year and quarter, the MS-DRG with its grouper version, patient disposition and length of stay, so it supports both AHRQ families simultaneously and is immediately available under a click-through agreement.

From 12,429,628 raw discharges we defined a single analysis cohort once, in a preparation step that every downstream configuration reads. We excluded records whose reported discharge quarter disagreed with the file quarter, patients under 18, and Major Diagnostic Category 14 (pregnancy and childbirth) and 15 (newborns). No record was excluded for a missing or invalid MS-DRG.

Secondary diagnoses 1 through 24 and their POA indicators were passed to ecsr10; the principal diagnosis was never passed. The only POA values present were Y, N, U, W and blank; 559 cells across the four years carried an invalid marker and were blanked.

One prespecified property of this cohort bounds one of the results that follow. The free Texas files end in 2019, so the applied cohort predates every Refined release it is scored under. This is immaterial to the POA and MS-DRG contrasts, but release choice affects an encounter through two mechanisms, the content of the code table and the ICD-10-CM version mapping that each release truncates at its own maximum, and only the first can act here: discharges from 2016 Q1 to 2019 Q4 resolve to ICD-10-CM versions 33 through 37, below the cap of every Refined release (v2021.1’s cap of 38 is the lowest). The earliest quarter at which the second mechanism can produce a release difference is 2021 Q4, when v2021.1’s cap first binds; between v2022.1 and later releases it is 2022 Q4. We therefore treat every release-contrast figure below as a lower bound on release sensitivity. The mechanism that does not contribute here is exercised instead by the multi-release validation dataset.

The applied study is reported according to the RECORD statement for observational studies using routinely collected health data [15] (S1 Checklist).

### Applied contrasts and outcomes

The identical predefined cohort was scored fourteen ways (Table 2). Eight configurations used the Refined family: the six releases v2021.1–v2026.1 with POA, and two POA counterfactuals at v2026.1 representing the two ways an analyst might handle a file without usable POA. *naiveY* overwrites every POA value to Y and leaves POA scoring on; it is the direct representation of a file that carries no POA at all, and the primary no-POA counterfactual. *poaoff* disables POA scoring and reproduces AHRQ’s POA-off macro setting exactly; it is an agreement check against SAS rather than an analytic recommendation, because AHRQ’s documentation states that the indices assume all 38 measures are coded. Six configurations used the beta family: the five releases with the MS-DRG screen applied, and one with the screen disabled to isolate its effect. For comparison across families, v2021.1’s two differently named measures were renamed to their v2022.1 spellings, which AHRQ’s change log records as a pure rename.

**Table 2.** The fourteen scoring configurations applied to the predefined cohort. All Refined configurations use the same diagnosis and POA columns; all beta configurations use the same diagnosis and MS-DRG columns. The reference configuration for every contrast reported in Results is Refined v2026.1 with POA.

| Configuration | Family | Release | Pre-existing-condition setting | Role |
| --- | --- | --- | --- | --- |
| v2021.1 | Refined | 2021.1 | POA indicators | release contrast |
| v2022.1 | Refined | 2022.1 | POA indicators | release contrast |
| v2023.1 | Refined | 2023.1 | POA indicators | release contrast |
| v2024.1 | Refined | 2024.1 | POA indicators | release contrast |
| v2025.1 | Refined | 2025.1 | POA indicators | release contrast |
| **v2026.1** | Refined | 2026.1 | POA indicators | **reference** |
| *naive Y* | Refined | 2026.1 | every POA value overwritten to Y, POA scoring enabled | primary no-POA counterfactual |
| *poaoff* | Refined | 2026.1 | POA scoring disabled (AHRQ’s POA-off macro setting) | counterfactual (SAS-equivalent specification) |
| beta v2016.2 | Beta | 2016.2 | 24 MS-DRG screens | release contrast |
| beta v2017.2 | Beta | 2017.2 | 24 MS-DRG screens | release contrast |
| beta v2018.1 | Beta | 2018.1 | 24 MS-DRG screens | release contrast |
| beta v2019.2 | Beta | 2019.2 | 24 MS-DRG screens | release contrast |
| beta v2020.1 | Beta | 2020.1 | 24 MS-DRG screens | beta baseline |
| beta, screen off | Beta | version-matched | MS-DRG screens disabled | sensitivity (isolates the screen) |

“Version-matched” means the beta release whose MS-DRG grouper vintage matches the grouper version recorded on the encounter; Texas regroups each calendar year with one grouper, so 2017, 2018 and 2019 map to beta v2017.2, v2018.1 and v2019.2, while calendar 2016 (grouper V33) has no matching beta release and v2020.1 has no matching year.

The *poaoff* configuration was scored directly on one quarter and derived for the remaining fifteen by copying the 20 POA-neutral measures and writing missing into the 18 POA-dependent ones; the scored and derived results were identical. The reference configuration throughout is Refined v2026.1 with POA. For each other configuration we report the proportion of encounters with at least one of the 38 (or 31, for beta) flags changed, the proportion whose mortality index changed, and the mean absolute index change. Agreement between the two families was quantified measure by measure with Cohen’s *κ* after mapping each beta measure to the Refined measure or measures covering the same construct.

### Statistical analysis

For the clinical-consequence analysis the outcome was in-hospital death (patient disposition “expired”). We drew one 70/30 train–test split stratified by discharge quarter and reused it for every configuration, so that the scoring implementation is the only component that varies. Models were logistic regressions of in-hospital death on age, sex and a comorbidity term, either the AHRQ mortality index or the 38 binary flags, all fitted on the same full training set. We report area under the receiver operating characteristic curve and the Brier score on the held-out 30% test set, following standard reporting practice for prediction models [23].

Because the models are fitted once and their predictions are then fixed, an interval on out-of-sample discrimination requires resampling only the test set, which we did with a nonparametric bootstrap of 2,000 replicates. Encounters within a hospital share coding practice, case mix and POA-reporting behavior, so the resampling unit was the hospital rather than the record: the 722 hospital codes present in the test set, plus the 12,650 encounters carrying the two masked provider IDs (999999 and 999998) that the Texas Department of State Health Services substitutes for small hospitals, which entered as singleton clusters because each masked ID pools many providers, giving 13,372 clusters in all. Dropping those encounters altogether moved no interval bound by more than 0.0013. Each replicate scored every configuration on the same drawn clusters, so differences between configurations are paired within replicate; we report the 2.5th and 97.5th percentiles. For comparison we repeated the procedure resampling records rather than hospitals. The intervals are conditional on the fitted models: they cover sampling of the test set, not uncertainty in the coefficients or in the split, a point we return to under Limitations.

All analyses used R 4.3.3 and ecsr10 on a single 12-thread workstation, with comorbidity() fed one quarter at a time to bound peak memory. A multicore scaling benchmark of comorbidity() on one quarter, with each core count measured in a separate R process and its flag matrix hashed so that a speedup could not change an answer unnoticed, is described in S2 Text.

### Ethics statement

The Texas Inpatient PUDF is a public-use file of de-identified discharge records, distributed by the Texas Department of State Health Services under a click-through data-use agreement that permits research use and forbids redistribution of records and any attempt at re-identification. The study used no identifiable private information and was not human subjects research; institutional review board review was therefore not sought. Only aggregate results are reported.

## Results

### Exact agreement with AHRQ across all supported releases

Across all three datasets and all eleven releases, 40,027,114 value-level comparisons produced zero differences (Table 3). This includes both the released POA-on configuration and the POA-disabled configuration, and both software families.

**Table 3.** Agreement between ecsr10 and the AHRQ SAS reference programs. Value columns are the comorbidity flags plus, where the release publishes them, the two index scores; cells compared excludes the encounter identifier used for row alignment. The beta comparison covers 29 columns rather than 31 because AHRQ’s program drops two hypertension components in favor of their union, which `ecsr10` also emits; the two are removed before comparison.

| Reference dataset | Family and releases | Encounters ×<br>releases | Value<br>columns | Cells compared | Differing<br>cells |
| --- | --- | --- | --- | --- | --- |
| Vendored AHRQ reference dataset | Refined, v2022.1–v2026.1 | 167,322 × 5 | 40 | 33,464,400 | 0 |
| Multi-release test dataset, POA on | Refined, v2021.1–v2026.1 | 11,954 × 6 | 38 or 40 | 2,845,052 | 0 |
| Multi-release test dataset, POA off | Refined, v2021.1–v2026.1 | 11,954 × 6 | 38 or 40 | 2,845,052 | 0 |
| Beta MS-DRG test dataset | Beta, v2016.2–v2020.1 | 6,018 × 5 | 29 | 872,610 | 0 |
| **Total** |  |  |  | **40,027,114** | **0** |

| Reference dataset | Family and releases | Encounters ×<br>releases | Value<br>columns | Cells compared | Differing<br>cells |
| --- | --- | --- | --- | --- | --- |

The multi-release test dataset is the dataset that discriminates releases. The same 11,954 encounters produce a different number of set flags under each of the six Refined releases, and both implementations produce the same six numbers. One synthetic encounter illustrates the mechanism: a heart-valve code (Q238) with POA = N and a discharge date in 2025 Q4 is POA-exempt under the ICD-10-CM version each of v2021.1–v2025.1 resolves that date to, but not under version 43, which v2026.1 alone reaches, so SAS and ecsr10 both set the flag under the first five releases and clear it under the sixth. Missing, invalid and out-of-range discharge dates are all handled by the SAS program’s final branch on both sides.

The beta configurations show the same pattern on the factor that varies there, the MS-DRG screen: disabling it on the beta test dataset changes 617 encounters. The beta versions are not all told apart by their totals, however: v2019.2 and v2020.1 each set 11,515 flags on this dataset yet disagree on 12 individual cells, which is why the beta comparison, like the Refined one, is made cell by cell.

The POA-disabled configuration deserves separate mention. AHRQ’s program initializes the 18 POA-dependent measures only inside its POA branch, so with POA disabled those measures reach the output dataset missing while their columns remain present. ecsr10 reproduces this and agrees with the SAS output on every one of the 478,160 compared cells at v2026.1 and on all six releases. Both index scores follow, because treating a missing flag as zero is arithmetically what the SAS summation in AHRQ’s program does when it skips a missing term; the consequence, which the output does not indicate, is that a POA-disabled index is computed over the 20 POA-neutral measures only and returns an ordinary number rather than a missing value. AHRQ’s user guide [11] is explicit that this is not the intended use: “the intended use of the weights to calculate the two comorbidity indices is specific to data that includes POA indicators” and “the indices depend on all 38 comorbidity measures being coded in the data”. The POA-disabled configurations reported below are therefore counterfactuals that quantify the effect of the choice, not a configuration we recommend.

### Third-party implementations diverged outside simple code mapping

Both packages we evaluated derive correct code tables, identical to ours at every release they implement, and then diverge from the reference in other processing steps; neither implements v2021.1 or any beta version (S1 Text). medicalcoder 0.9.0 completes every record, and its divergence is mainly a capability boundary: its scoring function accepts no discharge date, so one date-blind POA-exempt table stands in for eleven ICD-10-CM versions, which accounts for 86.2–91.0% of its differing cells at every release. The remainder, 131 cells over 59 encounters, is a de-duplication defect whose key omits the ICD code, so a code with missing POA can displace another code whose POA is Y. risk-compose 1.0.2 does not complete a run on this input: an undeclared intermediate measure aborts 31.7% of encounters, and neutralizing that exposes three further silent defects, in POA-exempt coverage, the cerebrovascular override and null handling. Outside AHRQ’s documented input requirements the four implementations are each lenient about a different subset of case, embedded decimals and padding, and agree on only 28 of 44 deliberately non-conformant records (S1 Text); none of this is a defect, but the same non-conformant file produces four different answers.

### Applied cohort

From 12,429,628 raw discharges, 11,523 were excluded because the reported discharge quarter disagreed with the file quarter, 2,253,845 for age under 18, and 1,579,016 for Major Diagnostic Category 14 (pregnancy and childbirth) or 15 (newborns), leaving a predefined cohort of 8,585,244 adult discharges from 726 reporting provider identifiers over 16 quarters (S1 Fig). Table 4 gives its characteristics. The cohort is predominantly older and acutely admitted (46.2% aged 65 or over, 60.5% emergency admissions, 49.8% with Medicare as expected primary payer), the population AHRQ derived both indices on.

**Table 4.**
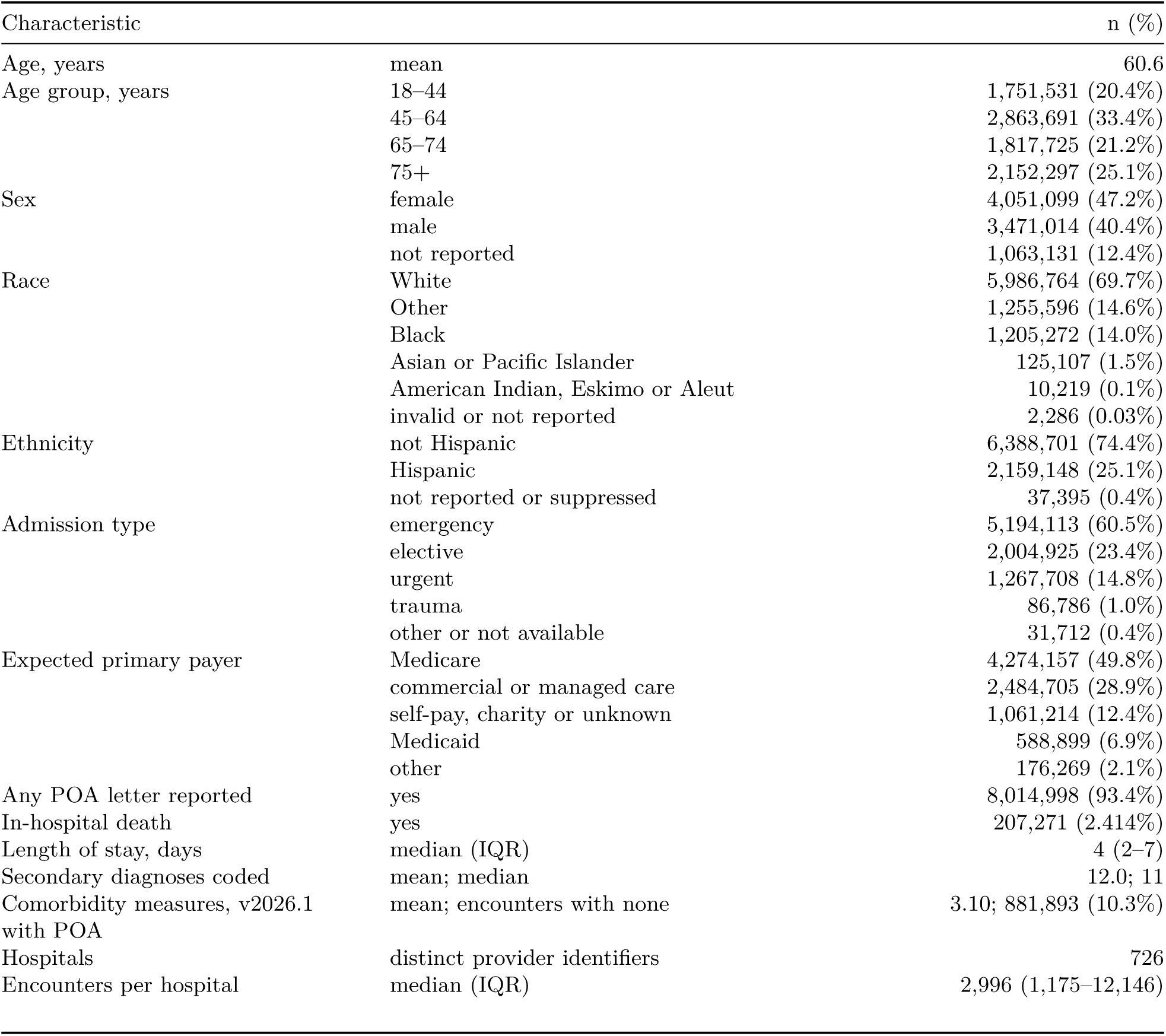
Characteristics of the Texas analysis cohort (n = 8,585,244). Values are n (%) unless stated. Age is grouped from the state’s banded age variable. Race, ethnicity, admission type and expected primary payer are decoded from the Texas PUDF data dictionary, with payer collapsed to five groups; the state recodes race to “Other” and blanks ethnicity for any race with fewer than ten discharges at a hospital. IQR, interquartile range.

The 12.4% of records with sex not reported is not missingness: Texas sets the sex code to “unknown” on any record carrying a human immunodeficiency virus (HIV)-, drug- or alcohol-related diagnosis, so sex-stratified results for the corresponding measures were excluded by prespecification.

Comorbidity prevalence at v2026.1 with POA was clinically plausible throughout, from uncomplicated hypertension at 38.9% and complicated hypertension at 23.2% down to carcinoma in situ at 0.026% (S1 Table), with a mean of 3.10 measures per encounter and 10.3% of encounters carrying none. Quarter-to-quarter prevalence was not flat (S2 Fig): between 2016 Q3 and 2016 Q4, when ICD-10-CM version 34 and the fiscal-year 2017 coding guidelines took effect, uncomplicated hypertension fell from 44.88% to 40.38% of encounters while complicated hypertension rose from 17.24% to 20.94%, and the two trends then continued gradually, reaching 36.70% and 26.18% by 2019 Q4. The same code table was applied to every quarter, so the step reflects coding practice, not the software.

### Release choice changed few flags but many index scores

Across 8,585,244 encounters and 38 measures, the AHRQ release had almost no effect on the comorbidity flags themselves (Table 5). Relative to v2026.1, v2023.1 through v2025.1 each differed on 13 encounters; v2022.1 differed on 310 (0.0036%); and v2021.1, the release that predates the ARTH-to-AUTOIMMUNE redefinition, differed on 32,922 (0.383%).

**Table 5.** Effect of each scoring choice on the comorbidity profile and the mortality index, relative to Refined v2026.1 scored with present-on-admission indicators (n = 8,585,244). v2021.1 publishes no index program, so index columns are not comparable for that configuration. ^a^Under *poaoff* every changed flag is a 1 in the reference configuration that becomes missing, never 0 to 1 or 1 to 0: disabling POA scoring leaves the 18 POA-gated measures unassigned, so this row counts encounters carrying at least one of those measures under v2026.1 with POA on. Missing is treated as 0 in the comparison.

| Configuration | Encounters with at least 1 flag changed | Mortality index changed | Mean absolute index change |
| --- | --- | --- | --- |
| v2021.1 | 32,922 (0.383%) | — | — |
| v2022.1 | 310 (0.0036%) | 42.44% | 0.558 |
| v2023.1 – v2025.1 (each) | 13 (0.00015%) | 0.00015% | <0.001 |
| *naiveY* (POA overwritten to Y) | 457,524 (5.33%) | 5.22% | 0.645 |
| *poaoff* (POA scoring disabled) | 4,867,045 (56.69%)<sup>a</sup> | 54.46% | 7.497 |

Only four ICD-10-CM codes drive the entire v2022.1-to-v2026.1 contrast on this cohort: O9081, O9902 and O9903, which mapped to blood-loss anemia through v2022.1 and were retired at v2023.1, and R939, which mapped to obesity through v2025.1 and was retired at v2026.1. In all, the v2022.1 and v2026.1 tables differ on 256 codes, 252 of them present only in v2026.1, added for codes that postdate the study window and do not occur in 8.6 million encounters. The larger v2021.1 gap arises from two unrelated changes at v2022.1: the removal of six gastrointestinal vascular codes from the peripheral vascular disease measure, and the redefinition of arthropathies as autoimmune conditions (Table 5). The second release mechanism, the truncated ICD-10-CM version mapping, contributed nothing here, as prespecified: discharges from 2016 Q1 to 2019 Q4 resolve to versions 33 through 37, below every release’s cap, so all six releases look up the same exemption list for every encounter (S2 Fig).

The index results differed sharply. AHRQ revised ten mortality weights once, between v2022.1 and v2023.1, and on this cohort that single change moved the mortality index for 42.44% of admissions, while the underlying flags were identical for all but 310 encounters (Table 5). The readmission weights have never been revised, and the readmission index was correspondingly identical across v2022.1–v2026.1. Reporting the release under which a comorbidity index was computed therefore matters even when the flags are provably unchanged.

### POA handling changed measure prevalence

The two POA configurations changed 5.33% and 56.69% of encounters, 14 times the largest release contrast (v2021.1) and more than a thousand times any release from v2022.1 onward (Table 5). Only the 18 POA-gated measures moved; the 20 POA-neutral measures remained unchanged, consistent with the specification (Fig 2). Overwriting every POA value to Y raised deficiency anemia by 1.46 percentage points, other neurological disorders by 1.04, coagulopathy by 1.03 and heart failure by 0.76.

**Fig 2.**
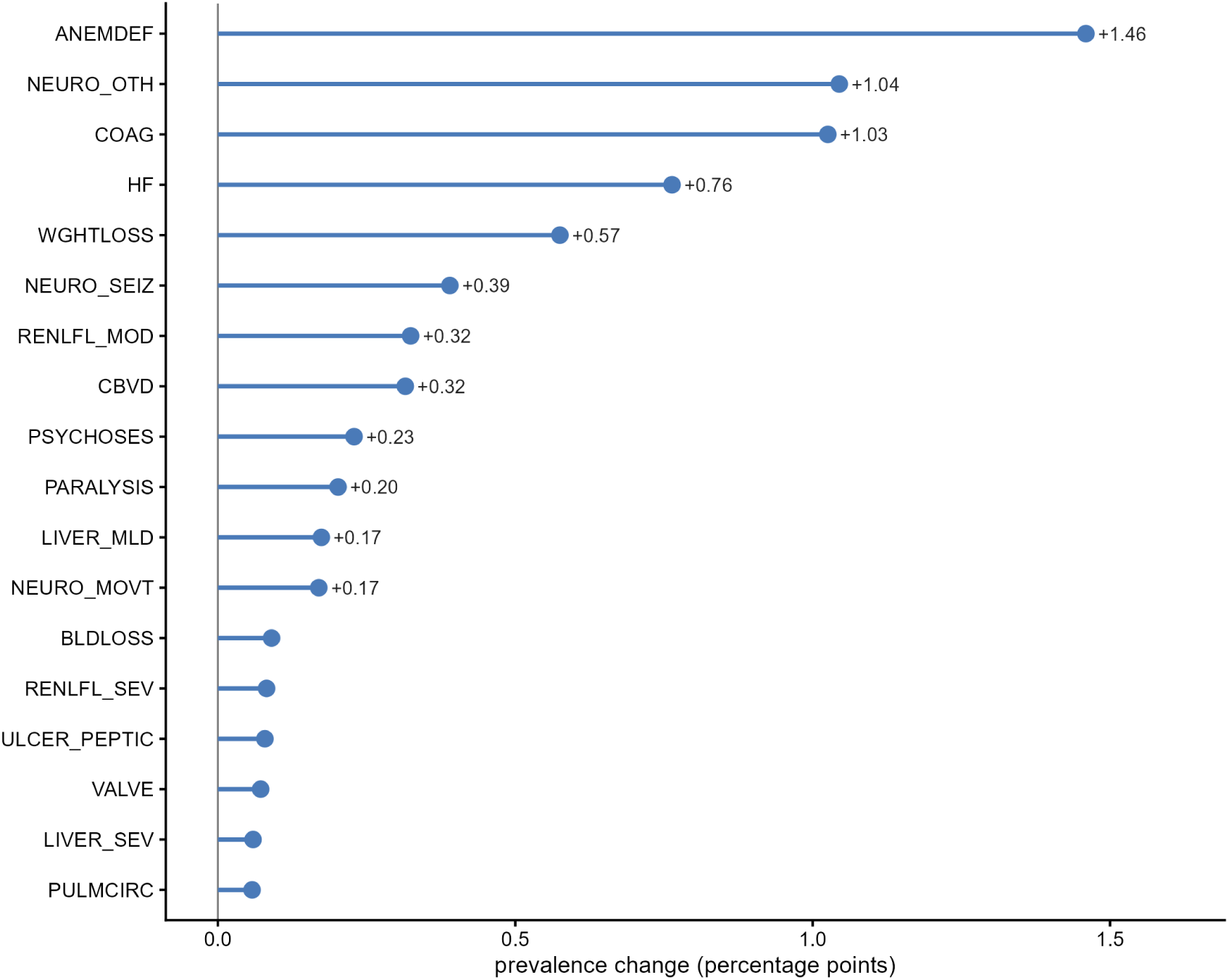
Prevalence change in the 18 present-on-admission-gated measures when present-on-admission indicators are ignored. Texas Inpatient PUDF 2016–2019. Percentage-point change in the prevalence of each of the 18 POA-gated measures at v2026.1 when every present-on-admission value is over-written to Y, computed from the v2026.1 and *naiveY* columns of S1 Table. The 20 POA-neutral measures are unchanged by construction and are not shown. Measures carry AHRQ’s abbreviations: ANEMDEF, deficiency anemias; NEURO_OTH, other neurological disorders; COAG, coagulopathy; HF, heart failure; WGHTLOSS, weight loss; NEURO_SEIZ, seizures and epilepsy; RENLFL_MOD, moderate renal failure; CBVD, cerebrovascular disease; PSYCHOSES, psychoses; PARALYSIS, paralysis; LIVER_MLD, mild liver disease; NEURO_MOVT, neurological disorders affecting movement; BLDLOSS, chronic blood-loss anemia; RENLFL_SEV, severe renal failure; ULCER_PEPTIC, peptic ulcer with bleeding; VALVE, valvular disease; LIVER_SEV, moderate-to-severe liver disease; PULMCIRC, pulmonary circulation disease.

The two POA conventions are not interchangeable. Disabling POA scoring leaves the 18 POA-gated measures missing, so every encounter carrying one of them in the reference configuration changes, and the cerebrovascular derivation, which sits inside the POA branch, does not run (4.725% prevalence in the reference configuration, 5.040% under *naiveY*, missing under *poaoff* ). Overwriting POA to Y keeps every derivation running and moves only the measures that POA gates, in one direction. The mortality index moves with the flags: it changed for 5.22% of encounters under *naiveY* and 54.46% under *poaoff* (Table 5, S3 Fig). The direct counterfactual for a file without POA is therefore to set the POA columns to Y and leave POA scoring enabled.

### POA handling changed mortality-model discrimination

On a single fixed 70/30 split stratified by discharge quarter, adding the AHRQ mortality index to age and sex raised out-of-sample discrimination from 0.6464 to 0.7885 (Table 6). Three findings follow.

**Table 6.** In-hospital mortality models, evaluated on the held-out 30% test set of a single 70/30 split (6,009,666 training and 2,575,578 test encounters, 62,332 test deaths). All models include age and sex and were fitted on the same training set. Confidence intervals are 2.5 and 97.5 percentiles of 2,000 bootstrap replicates resampling the test set clustered on hospitals (13,372 clusters), with the fitted models held fixed. AUC, area under the receiver operating characteristic curve; CI, confidence interval.

| Model | AUC (95% CI) | Brier score |
| --- | --- | --- |
| Age and sex only | 0.6464 (0.6381–0.6554) | 0.02346 |
| + mortality index, POA scoring disabled | 0.6898 (0.6835–0.6972) | 0.02333 |
| + mortality index, v2026.1 with POA | 0.7885 (0.7832–0.7945) | 0.02290 |
| + mortality index, v2022.1 with POA | 0.7886 (0.7833–0.7946) | 0.02289 |
| + mortality index, POA overwritten to Y | 0.8068 (0.8011–0.8129) | 0.02268 |
| + 38 flags, v2026.1 with POA | 0.7934 (0.7884–0.7989) | 0.02284 |
| + 38 flags, POA overwritten to Y | 0.8118 (0.8066–0.8174) | 0.02260 |

Differences between configurations are reported with 95% confidence intervals from a hospital-clustered bootstrap, paired within replicate (S4 Fig); a standard bootstrap that resamples individual records instead returns intervals up to 4.2 times narrower.

First, the single index recovers most of the discrimination available from all 38 flags. The mortality index alone reached an area under the curve (AUC) of 0.7885, where all 38 binary flags reached 0.7934, a gain of 0.0050 (95% confidence interval, CI, 0.0039–0.0061) for 37 additional covariates. Second, disabling POA is not a usable fallback for outcome modeling. Adding the POA-disabled index to age and sex raised the AUC from 0.6464 to 0.6898, where the POA-aware index raised it to 0.7885, a gain of 0.1421 (95% CI 0.1346–0.1497) against 0.0434. The POA-disabled index therefore recovers well under half of the discrimination the POA-aware index adds, because 18 of the 38 measures are missing by design and correctly contribute zero to the index. Third, and least intuitive, ignoring POA makes the model look better while changing what the covariate means. The POA-naive index reached 0.8068 and the POA-naive 38-flag model 0.8118, exceeding their POA-aware counterparts by 0.0183 in both cases (95% CI 0.0161–0.0204 and 0.0162–0.0205). This gain does not indicate a superior comorbidity measure: the model is instead supplied with in-hospital complications coded as though they were pre-existing conditions, and an analyst comparing hospitals on such a score is partly ranking them on their complication rates.

Paired resampling also puts the release axis on the same footing. The v2022.1 index out-discriminated the v2026.1 index by 0.00009 (95% CI 0.00001–0.00018; S4 Fig), a difference detectable only because it is paired and roughly two hundred times smaller than the POA effect.

### Hospital-level POA reporting was bimodal

Across the cohort’s 102,728,130 populated secondary-diagnosis positions, 23.8% carried no POA letter, but blanks were almost entirely confined to diagnoses exempt from POA reporting: 97.08% of AHRQ-mapped, POA-exempt positions were blank, against 3.52% of AHRQ-mapped, non-exempt positions (S2 Table). Because ecsr10 derives exemption from its own version-keyed table rather than trusting the reported letter, those blanks have no effect on scoring, and only 3.52% of the diagnoses that actually matter lacked a POA letter.

The residual risk is concentrated rather than diffuse. Some 6.64% of encounters carried no POA letter anywhere, and for those encounters 11 of the 18 POA-dependent measures cannot be set at all, while the other seven can be reached only through POA-exempt codes: deficiency anemia, moderate renal failure and coagulopathy all read 0.00%, against 21.31%, 11.31% and 6.93% where POA was reported, whereas cerebrovascular disease still reads 3.75% against 4.79% and severe renal failure 1.69% against 8.61%. This varies systematically by hospital rather than at random (Fig 3, S3 Table). Of 565 hospitals with at least 1,000 encounters, 70 reported POA on fewer than 1% of their discharges, while 195 reported it on 99% or more. At the non-reporting hospitals a POA-naive score adds a median of 0.72 comorbidities per encounter and as many as 3.06, where a hospital reporting on 99% or more gains a median of 0.04. The mortality index alone discriminates less well among encounters without any POA letter (area under the curve 0.640 against 0.773 where POA is reported; unadjusted, full cohort), but overwriting POA to Y leaves that gap essentially unchanged (0.658 against 0.794), so it reflects case mix rather than the missing letters: in-hospital mortality is 0.85% against 2.53%. Any hospital-comparison study on such a file must condition on POA reporting or exclude the non-reporting hospitals. Pooling them disadvantages the non-reporting hospitals themselves: their pre-existing disease is under-counted, so their expected mortality is understated and their observed-to-expected ratios are inflated relative to hospitals that report.

**Fig 3.**
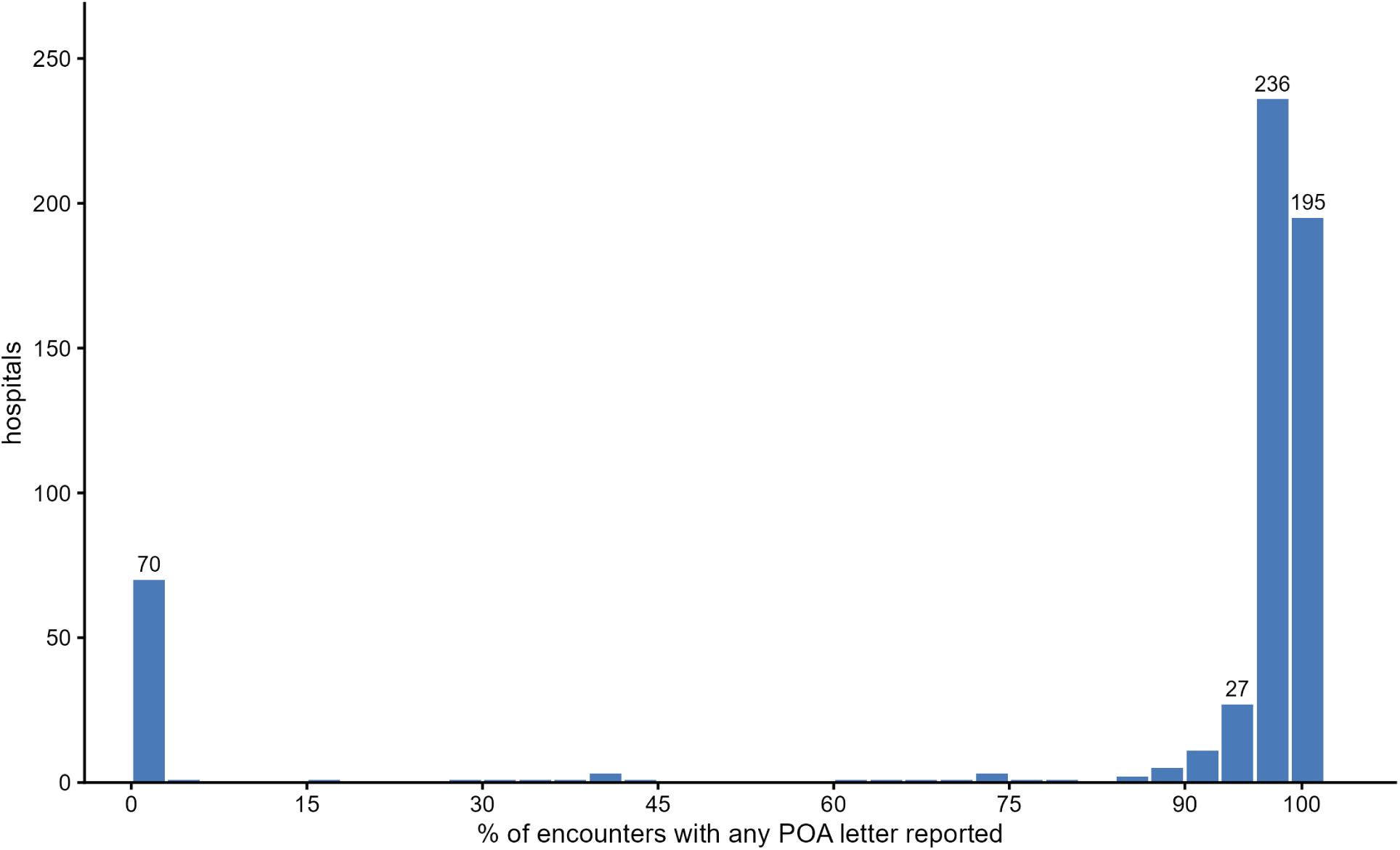
Distribution of present-on-admission reporting completeness across Texas hospitals with at least 1,000 encounters. Hospitals (n = 565) are binned by the percentage of their discharges carrying any POA letter in steps of 3 percentage points; bars are labeled where the bin holds at least 20 hospitals. Reporting is bimodal: 70 hospitals report on fewer than 1% of discharges (382,688 encounters) and 469 on 90% or more (8,017,661 encounters), leaving 26 spread across the range between. Per-hospital values are in S3 Table.

### Performance and availability

Scoring the full cohort under all fourteen configurations took a few hours on a single workstation, and one quarter of 524,253 encounters scored in 126 s on one core and 20 s on eight, with every core count returning an identical matrix of comorbidity flags (S2 Text). The web application scored both reference test datasets through the same pipeline the browser uses: its beta output reproduced AHRQ’s beta SAS output for every beta version, its Refined output passed the same SAS comparison script that gates the package, and a headless suite exercised the interface itself.

### The beta family and its MS-DRG screen

The beta family showed the same ordering on its own pre-existing-condition test (S3 Text). Scoring each encounter under its version-matched beta release with and without the MS-DRG exclusion screen, the screen suppressed at least one flag on 19.92% of encounters, a larger effect than any POA-handling choice in the Refined family except disabling POA altogether, and removed 39.8% of valve-disease flags (S3 Text). Beta release choice moved more than Refined release choice, changing at least one flag on 1.514% of encounters between v2016.2 and v2020.1 (S4 Table). The two families are not interchangeable: beta and Refined pulmonary circulation disorders share only 42% of their codes (*κ* = 0.10), and beta fluid and electrolyte disorders, at 31.2% the most common beta measure after hypertension, has no Refined counterpart at all (S3 Text).

## Discussion

We set out to ask which of the choices AHRQ’s Elixhauser Comorbidity Software leaves to the analyst actually change a result on real data, and to do so with an implementation whose fidelity to AHRQ’s SAS programs was established rather than assumed. The answer to the first question is that the analytic choice most consequential in practice is not the one the releases are designed around: present-on-admission handling moved far more than release choice did. The answer to the second is that exact equivalence is achievable and demonstrable: 40,027,114 value-level comparisons across eleven releases and two software families, with zero differences.

### Principal findings

Four results have direct implications for how comorbidity-adjusted analyses of administrative data are conducted and reported.

First, AHRQ’s Elixhauser software can be reproduced exactly outside SAS, across both families and all eleven releases, and the reproduction is checkable: ecsr10 agreed with AHRQ’s programs on every one of 40,027,114 compared cells, on test datasets constructed so that every release could be told apart. The two independent reimplementations scored on the same datasets did not, and their divergences lay in reshaping, date handling and the few derivations that are not simple lookups rather than in the code tables. An exactly validated implementation is what turns the remaining findings from arguments into measurements, because every difference between two scoring configurations is then a property of the configuration and not of the software.

Second, present-on-admission handling had the largest impact of any scoring choice. The two defensible conventions for a file without usable POA changed the comorbidity profile of 5.3% and 56.7% of admissions respectively. They are not approximations of one another: disabling POA scoring leaves 18 measures missing and skips the derivations that live inside the POA branch of AHRQ’s program, while overwriting POA to “present” keeps every derivation running. The direct counterfactual for a POA-free file is to set POA to “present” and leave POA scoring enabled, and the difference between that and the alternative is large enough to change a published prevalence. That diagnosis timing changes measured comorbidity is not new [12–14]; what these figures add is the size of the effect under each convention of the software most studies actually use.

Third, ignoring present-on-admission indicators improved apparent model performance while degrading the construct validity of the covariate. The POA-naive mortality index out-discriminated the POA-aware one by 0.0183 in area under the curve (95% CI 0.0161–0.0204, from a bootstrap clustered on hospitals), an improvement far larger than its own sampling variability that, taken at face value, would favor the wrong analysis. What the naive score adds is in-hospital complications counted as pre-existing disease. That is precisely the confounding that diagnosis-timing indicators were introduced to remove [12–14], and a form of information leakage that is easy to overlook when prediction models are built from routinely collected data [24]. Our hospital-level results translate this finding into an operational concern. POA reporting on this file is bimodal, with 70 of 565 hospitals reporting on under 1% of discharges and 195 reporting on 99% or more, and a POA-naive score adds a median of 0.72 comorbidities per encounter at the non-reporting hospitals against 0.04 where reporting is complete. A hospital-profiling analysis that pools these strata is partly ranking hospitals on their complication rates and partly on their documentation practices, a well-recognized hazard of administrative-data comparisons [13,25,26] that release-level software reproducibility alone does not address.

Fourth, the choice of release had minimal impact on the comorbidity flags but a substantial impact on the mortality index. Between v2022.1 and v2026.1 the Refined code tables differ on 256 ICD-10-CM codes, but only four occurred at all in 8.6 million Texas discharges, so at most 310 admissions (0.0036%) changed any flag; as prespecified, that figure is a lower bound, because the cohort predates every release compared. Over the same range, AHRQ’s one revision of ten mortality weights changed the mortality index for 42.4% of admissions. A study that reports “AHRQ Elixhauser comorbidity index” without naming the release has left a covariate undefined for 42.4% of its subjects, even though its comorbidity flags would have been reproducible from any release.

### What the validation contributes

Two features of the validation design generalize beyond this software. First, a test dataset must vary the factor being evaluated: a pipeline that scores one discharge date and never touches the few codes whose exempt status moves between versions cannot tell five of the six Refined releases apart, and a validation that reports agreement without demonstrating that its test dataset can disagree is reporting the dataset, not the software. Second, an exactly matching reference converts disagreement into adjudication: because ecsr10 matches AHRQ’s programs on every compared cell, every divergence another implementation shows is equally a divergence from AHRQ, which is what allowed us to classify medicalcoder 0.9.0’s date-blind POA-exempt table as a capability boundary and its de-duplication behavior as a defect by measurement rather than by argument. Both third-party packages derived correct code tables and diverged in reshaping, date handling and the few derivations that are not simple lookups, which is worth emphasizing because table content is what a reimplementation is usually checked on. For a user the practical question is narrower: for Refined releases with POA indicators, ecsr10 and medicalcoder 0.9.0 agree except where the discharge date matters, so a single-quarter file can use either, while a multi-year extract needs an implementation that resolves the ICD-10-CM version per record, and only ecsr10 covers v2021.1 or the beta family.

### Limitations

The applied cohort predates the releases whose differences we most wanted to measure. The free Texas files end in 2019, so v2023.1 through v2026.1 are separated by 13 encounters, and the ICD-version mechanism that makes releases differ in principle does not affect this cohort, because every discharge resolves to a version below every Refined release’s cap of 38 or higher. The earliest quarter at which that mechanism can separate releases is 2021 Q4 for v2021.1 and 2022 Q4 for v2022.1 against later releases. Everything we report about v2022.1 and later is therefore a lower bound on release sensitivity, and a replication on contemporary data, such as the HCUP State Inpatient Databases or Medicare MedPAR, would be expected to show larger flag differences, not smaller.

The confidence intervals we report on the mortality models bound one source of variability and not the others. They are computed by resampling the held-out test set, clustered on hospitals, with the fitted models held fixed, so they describe how much the reported discrimination would move on another draw of hospitals, not how much it would move under a different train–test split or a different fitted coefficient vector. We chose a single frozen split deliberately, because it is what makes the scoring implementation the only component that varies between arms. The price is that split-to-split variability is not quantified, and a repeated-split or temporally split replication would be a useful addition; the size of the effect argues that this is not where the conclusion is fragile, since the POA gap is roughly four times the width of its own confidence interval. The intervals are also a reminder that the record is the wrong sampling unit for this file: resampling records rather than hospitals returns intervals up to 4.2 times narrower.

The Texas PUDF carries no patient linkage, so the AHRQ readmission index is described here and never validated against an observed readmission, and mortality is in-hospital only; 30-day and post-discharge death are not observable. The file also sets the sex code to unknown on any record with an HIV-, drug- or alcohol-related diagnosis, which forced the exclusion of sex-stratified results for those measures. Data are from one state, and POA reporting behavior in particular is likely to differ elsewhere. The comparison with third-party implementations is an assessment of specific package versions (medicalcoder 0.9.0, risk-compose 1.0.2) on one test dataset; maintainers may have addressed these behaviors since, and the reproduction scripts are released so that the claims can be independently reproduced. The fully patched risk-compose configuration is a hybrid, that package’s logic over our reference tables, and shows that the algorithms converge once patched, not that the shipped package works.

Two boundaries of the software itself should be stated alongside the agreement result. ecsr10 has no analogue of the SAS programs’ per-record diagnosis count: it scores every diagnosis column it is given, so a file whose diagnosis count is shorter than its filled diagnosis list will differ from AHRQ’s programs run at their shipped defaults, and callers holding such a file must truncate the column list themselves. And AHRQ’s own scope statements bound interpretation rather than computation: the measures were refined on adult inpatient stays without explicitly addressing pediatric or outpatient care, the two indices were developed on adult, nonmaternal inpatient discharges, and their weights derive from 2018 data, with AHRQ noting that no analysis has been performed of the COVID-19 pandemic’s impact on them [11]. The Texas cohort is adult inpatient and contemporaneous with that derivation period, which is a strength for the release comparison but bounds how far the mortality-index result should be read forward into post-2020 data. These boundaries, together with the lack of any substitute for a missing POA indicator, apply to ecsr10 as to any implementation of this specification. Releases after v2026.1 will require a new code table and an extension of the date-to-version mapping; the software rejects an unrecognized release rather than defaulting silently. Finally, the web application has not been evaluated for usability; it is offered as a dissemination mechanism rather than as an evaluated interface.

### Implications and recommendations

Software identity is part of the method. On the evidence here, four items should be reported in any study using AHRQ Elixhauser comorbidities, and none of them is routinely reported today:

1. **The POA convention**: whether POA indicators were used, and if not, exactly which substitute rule was applied. This is the largest single source of variation we measured.
2. **POA completeness in the analytic file**, ideally by hospital, because a bimodal reporting distribution invalidates pooled hospital comparisons.
3. **The family and release**, for example “Refined v2026.1”, because the index weights changed once and the code tables change annually.
4. **The implementation and its version**, because two reimplementations of this same specification produce systematically different answers on dates and on encounters with partially missing POA.

The first three are algorithm-specification items of exactly the kind that reporting guidance for routinely collected health data already asks for [15], and the fourth is the software analogue. More broadly, this study is an argument for treating validation evidence as a citable research output accompanying the software rather than as internal quality control [19,21,27]; the test datasets, reference outputs, comparison script and adjudications are released with the package, so a reader can reproduce the claim rather than accept it.

## Conclusions

ecsr10 is an open-source R implementation of AHRQ’s Elixhauser Comorbidity Software that reproduces the reference programs exactly, across eleven releases, two software families and 40,027,114 value-level comparisons, where two independent reimplementations of the same specification do not. Applied to 8.6 million real discharges, it showed that present-on-admission handling changed more than half of all comorbidity profiles and inflated apparent model discrimination by counting inhospital complications as pre-existing disease, while release choice changed very few comorbidity flags yet moved the mortality index for 42.4% of admissions. Release, POA handling and the implementation used are analytic decisions with measurable consequences, and they belong in the methods section of every study that relies on them.

## Supporting information

**S1 Fig. Cohort size and in-hospital mortality of the Texas cohort by discharge quarter.** Texas Inpatient PUDF analysis cohort, 2016 Q1–2019 Q4.

**S2 Fig. ICD-10-CM code-set version resolved from discharge quarter across the study period.** The resolved version is identical under all six Refined releases.

**S3 Fig. Cumulative distribution of the AHRQ mortality index under three present-on-admission scoring conventions.** Texas cohort, Refined v2026.1.

**S4 Fig. Paired bootstrap differences in out-of-sample discrimination between mortality-model configurations, with hospital-clustered and record-level confidence intervals.** The six differences between mortality-model configurations reported in the Results, each taken within a bootstrap replicate so that the arms being compared share the same resampled hospitals. Wide bars are 95% confidence intervals from resampling hospitals; narrow bars are the intervals a bootstrap that resamples records instead returns. The two panels carry separate horizontal scales: what a comorbidity score adds over age and sex is two orders of magnitude larger than any difference between scoring configurations, and on one scale the latter would be indistinguishable from zero.

**S1 Table. Comorbidity measure prevalence, all configurations and strata.** Prevalence (%) of each of the 38 Refined measures in the predefined Texas cohort (n = 8,585,244) under each of the eight Refined scoring configurations, together with prevalence within the two present-on-admission reporting strata and their ratio. The configurations are the six AHRQ releases scored with POA and the two POA counterfactuals defined in Table 2 of the main text.

**S2 Table. Present-on-admission reporting by diagnosis type, 2016 Q1–2019 Q4.** The 102,728,130 populated secondary-diagnosis positions of the Texas cohort, split by whether the code is mapped by AHRQ Refined v2026.1 and whether it is POA-exempt, with the number and percentage carrying a blank POA indicator and the distribution of the reported letters. Exemption is decided per position against the POA-exempt list for the ICD-10-CM version its own discharge quarter resolves to, so the sixteen quarters contribute at versions 33 through 37 rather than at one shared version. Blanks are almost entirely confined to POA-exempt diagnoses (97.08% of AHRQ-mapped, POA-exempt positions, against 3.52% of AHRQ-mapped, non-exempt positions), and because ecsr10 derives exemption from its own version-keyed table those blanks have no effect on scoring.

**S3 Table. Present-on-admission reporting completeness by hospital.** One row per hospital with at least 1,000 encounters (n = 565). poa_rate is the percentage of that hospital’s discharges carrying any POA letter; poa_letters_per_dx the mean number of POA letters per populated secondary-diagnosis position. Index and measure columns give the hospital’s mean AHRQ mortality index and mean number of comorbidity measures under the reference configuration (v2026.1 with POA) and under the POA-naive configuration, with index_gap and como_gap their differences. death_pct is observed in-hospital mortality.

**S4 Table. Beta-family release drift against fixed and version-matched baselines.** Encounters and flag cells differing between each beta release and the baseline, on the full Texas cohort. level = "pooled" rows give totals per release; level = "by measure" rows break the same contrast down by comorbidity measure, with the measure name carried in the baselines column. basis distinguishes the fixed-baseline contrast from the version-matched one.

**S1 Text. Divergences of two independent reimplementations from the AHRQ SAS reference, and input-leniency behavior of four implementations.** Design and finding-level detail of the comparison of medicalcoder 0.9.0 and risk-compose 1.0.2 against AHRQ’s SAS programs on the multi-release test dataset, with the mechanism of each divergence and its attribution to a capability boundary or a defect, and the behavior of all four implementations on deliberately non-conformant input. Carries the finding-level divergence table (Table B) and the 44 input-leniency records (Table C).

**S2 Text. Multicore performance benchmark of comorbidity().** Design and result of the multicore scaling benchmark on one quarter of the Texas cohort, including the memory and superlinear-speedup behavior that motivates scoring large files in blocks, with the scaling curves drawn in Fig A.

**S3 Text. The beta family and its MS-DRG exclusion screen on the Texas cohort.** The effect of the beta family’s 24 MS-DRG exclusion screens on the Texas cohort, beta release drift, and the measure-by-measure comparison of the beta and Refined families. Carries the per-measure suppression figure (Fig A) and the beta-to-Refined crosswalk with code coverage and Cohen’s kappa (Table A).

**S1 Checklist. RECORD reporting checklist.** Completed REporting of studies Conducted using Observational Routinely-collected health Data (RECORD) checklist, the STROBE extension for routinely collected health data, with the manuscript section addressing each item.

## Author contributions

**Conceptualization:** Dong Pei. **Data curation:** Dong Pei. **Formal analysis:** Minh Tran, Dong Pei. **Investigation:** Minh Tran, Dong Pei. **Methodology:** Minh Tran, Dong Pei. **Software:** Minh Tran, Dong Pei. **Validation:** Dong Pei. **Visualization:** Minh Tran, Dong Pei. **Writing – original draft:** Minh Tran, Dong Pei. **Writing – review and editing:** Dong Pei.

## Data availability

The Texas Inpatient Public Use Data File is available from the Texas Department of State Health Services (https://www.dshs.texas.gov/center-health-statistics/texas-health-care-information-collection/download-and-purchase-data/texas-inpatient-public-use-data-file-pudf) under a data-use agreement that forbids redistribution of record-level data, so the record-level data cannot be shared by the authors. All aggregate result tables underlying the reported findings are provided as Supporting Information files (S1–S4 Table).

## Code availability

ecsr10 is available at https://github.com/dpei/ecsr10 under an MIT license. The validation pipelines, the applied-analysis pipeline and the web application are released at https://github.com/dpei/ecsr10_simulation; the web application is also hosted for direct use at https://slicer.shinyapps.io/ecsr10/.

## AI tool usage declaration

During the preparation of this work, we used Claude (Anthropic) for refining scientific writing and code generation. After using this tool, we reviewed and edited the content as needed and take full responsibility for the content of the publication.

